# Quantifying the impact of hospital catchment area definitions on hospital admissions forecasts: COVID-19 in England, September 2020 - April 2021

**DOI:** 10.1101/2023.07.12.23292451

**Authors:** Sophie Meakin, Sebastian Funk

## Abstract

**Background:** Defining healthcare facility catchment areas is a key step in predicting future healthcare demand in epidemic settings. Forecasts of hospitalisations can be informed by leading indicators measured at the community level. However, this relies on the definition of so-called catchment areas, or the geographies whose populations make up the patients admitted to a given hospital, and which are often not well-defined. Little work has been done to quantify the impact of hospital catchment area definitions on healthcare demand forecasting.

**Methods:** We made forecasts of Trust-level hospital admissions using a scaled convolution of local cases (as defined by the hospital catchment area) and a delay distribution. Hospital catchment area definitions were derived from either simple heuristics (in which people are admitted to their nearest hospital or any nearby hospital) or historical admissions data (all emergency or elective admissions in 2019, or COVID-19 admissions), plus a marginal baseline definition based on the distribution of all hospital admissions. We evaluated predictive performance using each hospital catchment area definition using the Weighted Interval Score (WIS) and considered how this changed by the length of the predictive horizon, the date on which the forecast was made, and by location. We also considered the change, if any, on the relative performance of each definition in retrospective vs. real-time settings, or at different spatial scales.

**Results:** The choice of hospital catchment area definition affected the accuracy of hospital admission forecasts. The definition based on COVID-19 admissions data resulted in the most accurate forecasts at both a 7- and 14-day horizon, and was one of the top two best-performing definitions across forecast dates and locations. The “nearby” heuristic also performed well, but less consistently than the COVID-19 data definition. The marginal distribution baseline, which did not include any spatial information, was the lowest-ranked definition. The relative performance of the definitions was larger when using case forecasts compared to future observed cases. All results were consistent across spatial scales of the catchment area definitions.

**Conclusions:** Using catchment area definitions derived from context-specific data can improve local-level hospital admissions forecasts. Where context-specific data is not available, using catchment areas defined by carefully-chosen heuristics are a sufficiently-good substitute. There is clear value in understanding what drives local admissions patterns, and further research is needed to understand the impact of different catchment area definitions on forecast performance where case trends are more heterogeneous.

## Introduction

Short-term forecasts were widely used during the COVID-19 pandemic to support public health policy through planning and resource allocation. During 2020 and early 2021, it was an ongoing concern that demand for hospital care would exceed capacity.

Defining hospital catchment areas serves a number of purposes, primarily for healthcare administration. First, to estimate the size of each catchment population - the number of people using, or who may use, the health services - and the catchment population’s demographics. This facilitates planning, such as allocating budgets and determining staffing and other resource needs. Second, to identify regions with under- or overprovision of hospital services. Finally, to calculate admissions rates per capita or estimate vaccination coverage, using the catchment population size as the population denominator.

This last application is also highly relevant for modelling of hospital admissions, including short-term forecasting. In England, local-level COVID-19 cases and hospital admissions are reported on different scales: cases are reported by local authority (a small subnational administrative unit), and admissions by NHS Hospital Trusts (comprising a small number of hospitals providing care to a small geographical region or for a specialised function). Previous evaluation of local-level COVID-19 hospital admissions forecasts have shown that using cases as a leading predictor can improve forecast accuracy [1]; however, to use “local” cases as a predictor of hospital admissions requires the forecaster to define “local”, that is, to define each hospital’s catchment area.

Hospital catchment areas in England are not well-defined, since prospective patients have a right to choose where to seek health care [2]. According to the Office for Health Improvement and Disparities, NHS Trusts do not have geographically defined boundaries for service access, nor a complete and up-to-date list of registered patients [3]. Any proposed catchment area definition should aim to capture a significant proportion of the hospital’s activity, reflect physical barriers to access (travel distance or time), differ by speciality and increase with hospital size [4].

Hospital catchment areas are estimated using geographically-linked admissions data, or simple heuristics. If data on historical admissions is available, then this data could be used with a model to define the catchment areas. Two simple models are frequently cited: first-past-the-post, where geographical areas are assigned to the single hospital to which the majority of patients sought care, and proportional flow, where populations are split between hospitals according to the proportion of admissions to each hospital (Jones et al. 2011). The UK Office for Health Improvement and Disparities uses the proportional flow model [3]. Both the first-past-the-post and proportional flow models assume no future change in hospital admissions patterns. Gravity models [5,6], and other more complex regression models [7], are also used but require yet more data on population demand, hospital capacity, and travel distances or times between patients’ homes and hospitals. Using more complex models may also reduce understanding, interpretability, and uptake for stakeholders unfamiliar with modelling methodology.

Geographically-linked hospital admissions data may not always be readily available [8]. In these scenarios, heuristics can be used instead. The simplest assigns geographical areas to their nearest hospital using an adapted Voronoi decomposition (also known as a Thiessen decomposition) which conforms to administrative boundaries; this produces non-overlapping catchment areas where prospective patients are assumed not to have any choice in service provider. However, numerous studies in a wide range of settings have shown that patients often choose to travel further to seek alternative facilities [9–11]. Alternatively, catchment areas can be defined as any geographical region within a fixed distance [12] or, where appropriate and available, travel time [8,13]. Other algorithmic approaches include K-means clustering [4] and label propagation [14].

Estimated catchment areas and populations have been used to predict changes in emergency admissions in response to changes to hospital capacity in London [5], and to estimate the cumulative rate of confirmed malaria cases for six healthcare facilities in Uganda [12], amongst others [13]. Only a few studies considered the sensitivity of their results to the catchment area definition used [12], despite acknowledging that these areas were not well-defined. To our knowledge, no forecasting studies have assessed the sensitivity of quantitative forecasting performance to the choice of hospital catchment area definitions.

In this study, we evaluated the impact of using different hospital catchment area definitions on the accuracy of local-level hospital admissions forecasts, using COVID-19 during September 2020 - April 2021 as a case study. We used a scaled convolution of local cases (as defined by each of the catchment area definitions) and a delay distribution as the forecasting model. We showed previously this model makes accurate forecasts of local-level COVID-19 hospital admissions, under the condition that we can make consistently good case forecasts [1]. We measured and compared the performance of the resulting forecasts, and summarised average performance by forecast horizon, forecast date and location. We considered both a retrospective scenario, where future cases are assumed to be known in order to provide a best case for the forecast performance, and a real-time scenario, where future cases are not known and a case forecast used instead. We also explored whether there was any change in our main conclusions when using catchment areas defined at different spatial scales by comparing forecasting performance using catchment areas definitions by lower- and upper-tier local authority.

## Methods

### Data

#### Trust-level COVID-19 hospital admissions

A confirmed COVID-19 hospital patient is any patient admitted who has recently (in the last 14 days) tested positive for COVID-19 following a polymerase chain reaction (PCR) test, including both new admissions with a known test result and inpatient tests. Data on daily Trust-level COVID-19 hospital activity, including COVID-19 hospital admissions, COVID-19 and non-COVID-19 bed occupancy, are published weekly by NHS England and were accessed via the *covid19.nhs.data* R package [15].

#### LTLA-level COVID-19 cases

A confirmed COVID-19 case in England is defined as an individual with at least one confirmed positive test from a PCR, rapid lateral flow tests or loop-mediated isothermal amplification (LAMP) test. Positive rapid lateral flow test results can be confirmed with PCR tests taken within 72 hours; if the PCR test results are negative, these are not reported as cases. Aggregated data by lower-tier local authority (LTLA) are published daily on the UK Government dashboard [16] and reported totals include both pillar 1 (tests in healthcare settings and for health and care workers) and pillar 2 (community) tests. We use data published by specimen date, the date on which the test is taken.

#### Hospital catchment area definitions

Since COVID-19 cases and hospital admissions are reported on different scales (local authority and NHS Trust, respectively), we need to define each Trust’s catchment area to estimate COVID-19 cases “local” to each Trust.

We considered six definitions of hospital catchment areas for Acute NHS Trusts in England. The catchment area definitions are based on heuristics about where people may seek hospital care, or derived from data on historical hospital admissions. We defined a Trust’s catchment area with respect to local authority (UTLA or LTLA) boundaries, and for each local authority-Trust pair assigned a weight, p, that represented the expected proportion of people/admissions from that local authority that would seek hospital care at the given Trust. Catchment area definitions are defined to ensure that each local authority is matched to at least one Trust, and vice versa; consequently, the catchment area boundaries are overlapping, since each local authority can be mapped to more than one Trust.

The six catchment area definitions are as follows:

##### Marginal distribution (baseline)

Patients from all local authorities go to Trusts according to the marginal distribution of hospital admissions June 2020 - May 2021. For example, if Trust X admitted 10% of all COVID-19 patients in this time period, then the marginal distribution assigns a weight of 10% between all local authorities and Trust X.

##### Nearest hospital

Patients from each local authority go to any Trust within the local authority with equal probability, or to their nearest Trust with probability 1 if there are no Trusts in the local authority. Distance between local authorities and Trusts is measured as the Euclidean distance between the population-weighted centre of the local authority (based on shapefiles as of December 2021 [17,18]) and the main site of the Trust [18,19].

##### Nearby hospitals

Patients from each local authority go to any Trust within a 40 km radius from the population-weighted centre of the local authority. The value of 40 km was chosen to be larger than the maximum minimum local authority-Trust distance for any local authority; a lower threshold would result in a catchment area definition for which some local authorities were not mapped to any Trusts.

##### Emergency hospital admissions, 2019

Defined by the distribution of all emergency hospital admissions from UTLA to Trust in England during 2019 (January through December) [3].

##### Elective hospital admissions, 2019

Defined by the distribution of all elective (non-emergency and pre-planned) hospital admissions from UTLA to Trust in England during 2019 (January through December) [3].

##### COVID-19 hospital admissions, June 2020 through May 2021

Defined by the distribution of COVID-19 hospital admissions from UTLA to Trust in England during June 2020 - May 2021. Confirmed COVID-19 cases were linked to hospital admissions by case/patient ID [15]. For each case we keep at most one admission, namely the first admission only from all admissions where (i) the admission date was earlier than the test specimen date plus 28 days, and (ii) the discharge date was after the test specimen date.

### Comparison of catchment area definitions

We compared the six hospital catchment area definitions to understand where and how the definitions differed from one another: any difference in forecast performance under the different catchment area definitions will be contingent on differences in the definitions themselves, and on differences between trends in COVID-19 cases between local authorities within each catchment area. We compared the catchment area definitions using a number of summary statistics, as well as quantifying how much two catchment area definitions overlap. We also quantified the similarity of COVID-19 case time series between local authorities within Trusts’ catchment areas.

#### Descriptive statistics

We summarised and compared the following Trust-level properties of the hospital catchment area definitions. First, the distribution of catchment area weights. Second, the number of local authorities under each catchment area definition for which the assigned weight is more than x%, that is, for which at least 10% of the local authority population are assigned to that Trust; we consider x = 0%, 1%, and 10%. Third, the median distance from each Trust to any local authority for which the assigned weight is more than x%; again, we consider x = 0%, 1%, and 10%. Finally, we calculate the proportion of a catchment area’s total weight that is from either the nearest, or nearby (< 40 km), local authority or authorities.

#### Quantifying the similarity of catchment area definitions

To summarise and quantify the similarity between pairs of catchment area definitions for a given Trust, we defined an overlap-similarity metric. The *asymmetric overlap-similarity* between catchment area definitions *X* and *Y* relative to definition *X* is defined as the proportion of the definition *X* that is contained in the overlap with *Y*. The *overlap-similarity* between *X* and *Y* is then simply the minimum of the asymmetric overlap-similarity relative to each of *X* and *Y*. See the Supplementary Information Section 3.2.2, for details of these definitions.

By construction, the overlap-similarity is equal to zero if, and only if, the two catchment area definitions do not include any of the same local authorities. The asymmetric overlap-similarity between *X* and *Y*, relative to *X*, is equal to 1 if, and only if, the weights assigned by *X* are less than or equal to the weights assigned by *Y* for all local authorities. The overlap-similarity metric is equal to 1 if, and only if, the two definitions assign exactly equal weights to all local authorities. A higher overlap-similarity score indicates that *X* and *Y* are more similar, that is, the weights assigned to each local authority are similar for definitions *X* and *Y*.

#### Quantifying the similarity of COVID-19 case time series

We expect that any difference between forecasts made using different catchment area definitions will require some heterogeneity in the trends of reported COVID-19 cases in the local authorities in the catchment area - if nationwide trends are aligned, the definition of catchment areas should not matter. To this end, we used the Pearson correlation coefficient to quantify the similarity between COVID-19 case time series between local authorities. First, for each forecast date and pair of local authorities, we calculated the Pearson correlation between the time series of COVID-19 cases in each local authority from 21 days before until 14 days after the forecast date. Then for each forecast date we calculated two values. First for each local authority the median correlation coefficient with all other local authorities. Second for each Trust the median correlation coefficient between all local authorities for which any catchment area definition assigns a weight of more than 10%. The threshold value of 10% is arbitrarily chosen, but such that only the main local authorities in each catchment area were included.

### Admissions forecasting model

We made forecasts of hospital admissions using a convolution of estimated Trust-level COVID-19 cases with the delay from report to admission [1,20], henceforth referred to as the case-convolution model, for brevity. Full details of this case-convolution model are in the Supplementary Information Section 1.2. We showed previously that this model makes accurate forecasts of local-level COVID-19 hospital admissions: it clearly outperformed simple trend-led time series models, especially at longer forecast horizons, albeit under the condition that we were able to make good case forecasts [1].

To forecast admissions, the case-convolution model takes COVID-19 cases after the forecast date as an input. Here, we considered two scenarios: retrospective and real-time. In the retrospective scenario, we use future observed cases with no uncertainty, and so this represents a best-case scenario in which any difference in forecast performance is as a direct result of using the different catchment area definitions. Under this scenario, we evaluated forecast performance for both lower- and upper-tier local authority definitions of catchment areas.

In the real-time scenario, we used forecasts of cases that were made via estimates and forecasts of the real-time reproduction number, Rt, accounting for uncertainty in the delay distributions and day-of-the-week effect, produced and published daily [20,21]. A summary of this is given in the Supplementary Information Section 1.1 [1,20–23]. Under this scenario, we evaluate real-time forecast performance, as well as assess how forecast performance changes when uncertainty in future cases is included. As case forecasts were only made at the UTLA level, we evaluated forecast performance in the real-time scenario for the upper-tier local authority definition of catchment areas only.

### Forecast evaluation

We assessed different aspects of forecast performance using standard metrics, described below. We summarised forecast performance against these metrics by forecast horizon, by forecast date (the date on which each forecast is made, 15 total), and by location (acute NHS Trust, 138 total).

#### Calibration

Calibration measures models’ ability to correctly quantify predictive uncertainty. We assessed calibration by calculating the empirical coverage: for a given forecast horizon and prediction interval width, the empirical coverage of a model is the proportion of forecast targets (here, across all dates and locations) for which the prediction interval contains the true value. We calculated the empirical coverage for the 50% and 90% prediction intervals. A perfectly-calibrated model has empirical coverage equal to the width of the nominal prediction interval.

#### Probabilistic forecast error

We measured probabilistic forecast accuracy with the weighted interval score (WIS), a proper scoring rule, a rule for which a forecaster is incentivised to give their honest forecast to obtain the best score [24]. The WIS comprises a weighted sum of interval scores for quantile forecasts of increasing widths, measuring both the sharpness (the width of the forecast interval) and accuracy (whether the interval contains the true value) of each quantile forecast. By combining these interval scores, the probabilistic forecast accuracy of the full forecast distribution is summarised in a single value. The relative weighted interval score (rWIS) is an adjusted WIS value that allows us to compare the performance of different definitions over forecast dates or between Trusts, both of which vary in the number of admissions.

### Analysis code

Analyses in this paper use the following packages developed during the COVID-19 pandemic: *covid19.nhs.data* (0.1.0) [15], *EpiNow2* (1.3.3.8) [20], and *scoringutils* (0.1.7.2) [25,26]. Fully reproducible code is available at https://github.com/epiforecasts/hospitalcatchment-forecast and https://github.com/epiforecasts/hospitalcatchment.utils.

## Results

### Comparison of catchment area definitions

#### Catchment area definition descriptive statistics and overlap-similarity

A visual inspection reveals some clear differences between the hospital catchment area definitions (Figure 1); these differences were more evident from the descriptive summary statistics (supplementary Figure S3 - 5). The marginal distribution definition is clearly very different to all other definitions, since it includes all local authorities in the catchment area for every Trust; we do not discuss it further here. The other major differences were between the “nearest” and “nearby” heuristic definitions, and between the heuristic definitions and the three data-derived definitions.

**Figure 1:**
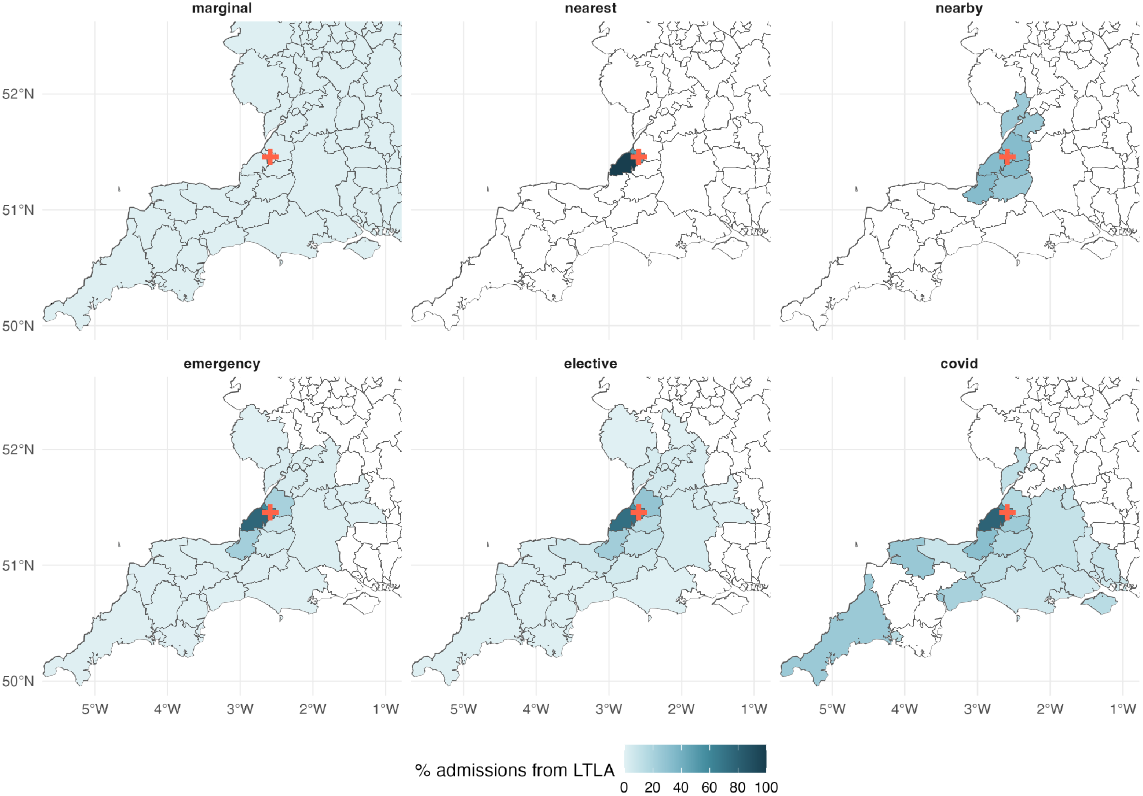
Comparison of six LTLA-level hospital catchment area definitions for University Hospitals Bristol And Weston NHS Foundation Trust. The hospital catchment areas are defined as follows: by the marginal distribution of hospital admissions to Trusts June 2020 - May 2021 (“marginal”); the nearest Trust for each LTLA (“nearest”); any Trust within 40 km of the LTLA (“nearby”); by the distribution of emergency, or elective, hospital admissions in 2019 (“emergency” and “elective”, respectively); and by the distribution of COVID-19 hospital admissions June 2020 - May 2021 (“covid”). In each panel, the colour denotes the proportion of all patients from that LTLA that are admitted to University Hospitals Bristol And Weston: darker colours indicate a higher proportion, and white indicates zero admissions. The Trust’s main site is marked by a red cross.

First, we compared the nearest and nearby heuristics. The characteristics of the nearby heuristic were very different for Trusts inside versus outside the London NHS region. In London, the distribution of weights was very homogeneous (median 0.037, IQR [0.037, 0.04]), each Trust had on average 38 UTLAs in its catchment area (IQR [38, 38]), and the median distance between Trusts and UTLAs was 14.4 km (IQR [13.4, 17.9]). This is because local authorities in London (boroughs) are small (all less than 10 km in diameter) and so the 40 km radius includes most other boroughs as well as other nearby local authorities. Outside of London, the distribution of weights was more heterogeneous, as was the median number of UTLAs per catchment area (median 7, IQR [3, 11]) and the median distance was higher (median 23.1 km, IQR [18.8, 29.5]). In comparison to the “nearby” heuristic, the nearest heuristic was very homogeneous and, by construction, constrained to a much smaller geographic area. The majority of Trusts (98/138) had a single UTLA in its catchment area (median 1, IQR [1, 2]). The median distance between Trusts and their nearest UTLA is very small (median 5.4 km and IQR [2.8, 14.7]; supplementary Figure S5A).

In contrast to the simple heuristics, the distributions of weights for the three definitions derived from admissions data were more heterogeneous (Figure 1 and supplementary Figure S3). More specifically, we found that the weight assigned to local authorities for a given Trust decreased as the distance between local authority and Trust increased. Compared to the heuristic definitions, the emergency, elective and COVID-19 admissions definitions shared many similarities. The number of local authorities in a Trust’s catchment area was comparable across the three definitions (median of 4, 5 and 3 UTLAs for the emergency, elective, and COVID-19 admissions data definitions, respectively, for a threshold of x = 1%; supplementary Figure S4, second column), as was the average distance (median 17.0, 18.2 and 14.3 km for the emergency, elective, and COVID-19 admissions data definitions, respectively, using a weight threshold of x = 1%; supplementary Figure S5A, second column). A large proportion of Trusts’ emergency, elective and COVID-19 admissions catchment areas are from the nearest local authority (median 76.5%, 72.4% and 83.1%, respectively; supplementary Figure S5B). Furthermore, virtually all of Trusts emergency, elective and COVID-19 catchment areas were from nearby local authorities (within 40 km) (median 99.3%, 97.8% and 100%, respectively; supplementary Figure S5C). By contrast, only 15.3% of the “nearby” heuristic definition was from the nearest local authority, since all local authorities within a Trust’s 40 km radius were assigned the same weight, irrespective of distance.

According to the overlap-similarity metric, the emergency and elective catchment area definitions were most similar (median overlap-similarity 0.84; supplementary Figure S6A), and both were also similar to the COVID-19 definition (0.74 and 0.77 median overlap-similarity with the emergency and elective definitions, respectively; supplementary Figure S6A). Moreover, the median asymmetric overlap-similarity relative to the COVID-19 definition was 1 and 0.99 for the emergency and elective admissions definitions, respectively (supplementary Figure S6B), that is, for more than half the Trusts all local authority weights assigned by the COVID-19 definition were less than or equal to the weights assigned by either the emergency or elective definitions.

As expected, the marginal distribution definition was very dissimilar to all other definitions, with a median overlap-similarity ≤ 0.05 with all other definitions (supplementary Figure S6A), and the majority of asymmetric overlap-similarity values < 20% (supplementary Figure S6C). The nearby heuristic was also dissimilar to the other definitions on average (median overlap-similarity < 0.3; supplementary Figure S6A), although individual asymmetric overlap-similarity values varied considerably from one Trust to another for all definitions except the marginal distribution (supplementary Figure S6C).

#### Similarity of COVID-19 case time series within catchment areas

Although trends in local COVID-19 cases often varied across England, cases in neighbouring local authorities were generally strongly correlated with each other (supplementary Figure S7). The median pairwise correlation by LTLA (averaged across the correlation with all other LTLAs in England) varied substantially, with some notable dates and locations where the median value was negative (supplementary Figure S7A). For example, the median correlation in Liverpool in mid-October and early-November 2020 was negative: while cases in most local authorities were rising, they were decreasing in Liverpool and nearby local authorities due to local restrictions on social distancing [27]. Another example: the median correlation in Medway (a mainly rural local authority in South East England) in the second half of November and early December 2020 was negative: cases in Medway were rising as the Alpha variant emerged, while cases were stable or declining in most local authorities following the second national lockdown (05 November - 02 December 2020) and additional earlier restrictions on social distancing.

In contrast, the number of cases reported by local authorities within the same catchment area were usually strongly correlated with each other (median correlation coefficient > 0.5; Figure S7B), especially October 2020 through February 2021. For example, in Liverpool NHS Foundation Trust, the median correlation between the main LTLAs in its catchment area (Liverpool, Knowsley, Sefton and West Lancashire) was above 0.7 throughout October and November 2020, despite the negative correlation nationally. Similarly, the correlation between the main LTLAs in the catchment area of Medway NHS Foundation Trust (Medway and Swale) was above 0.5.

### Forecasts comparison

Despite some clear differences between the six catchment area definitions, the median forecasts under each definition were, on average, strongly positively correlated with each other. This was likely as a result of high correlation between reported COVID-19 cases within the majority of catchment areas during the evaluation period. The median correlation coefficient (across all locations and dates) was above 0.8 for any pair of definitions (Figure S8A), and was especially high during October 2020, and December 2020 through February 2021 (Figure S8B). However, forecasts made under different catchment area definitions were less strongly correlated during other time periods. Notably, the median correlation coefficient (across all locations) between forecasts made on 29 November 2020 using the marginal distribution definition and any other definition was less than 25% (supplementary Figure S8B). This is likely due to the emergence of the Alpha variant in London and Kent in South East England and subsequent rise in cases following a period of varying local restrictions (the “tier system”) in the North of England and a month-long national lockdown.

Since the marginal distribution definition is not a local definition (the catchment area is the same for all Trusts), then it is unsurprising that in this very localised context that it leads to very different median forecasts than the other definitions. A visual inspection of the forecasts shows example forecast dates and locations for which the forecasts are meaningfully different (for example, Mid And South Essex NHS Foundation Trust on 13 December 2020: Figure 2).

**Figure 2:**
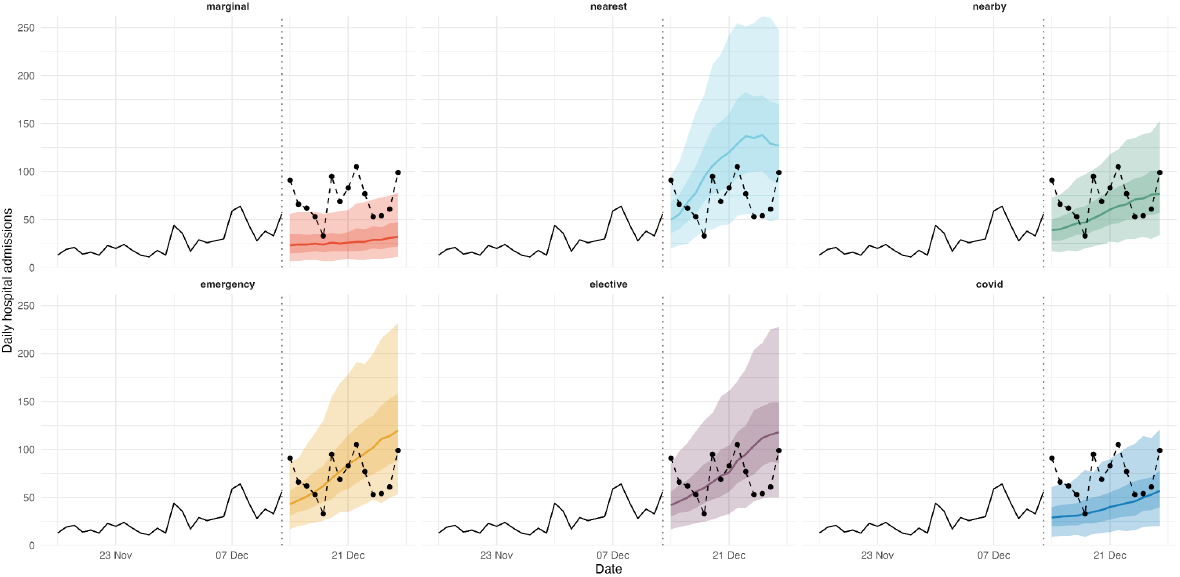
Example of retrospective forecasts made 13 December 2020 for Mid And South Essex NHS Foundation Trust. These forecasts are made based on UTLA-level catchment area definitions and using future observed cases. Shown are median forecasts (line), and 50% and 90% quantile forecasts (dark and light ribbon, respectively). The black solid line shows admissions observed up to the forecast date (13 December, marked by a vertical dotted line), while the black dashed line and points show realised admissions, for reference.

### Forecast evaluation

#### Calibration

All catchment area definitions resulted in forecasts that overestimated the uncertainty (Figure S9): for nominal coverage of 50%, the empirical coverage of all definitions was 60 - 70%, and for nominal coverage of 90% the empirical coverage was 90 - 95%. The difference between nominal and empirical coverage decreased, albeit not substantially, at longer forecast horizons. For example, for a nominal coverage of 50%, empirical coverage of all definitions was in the range 68 - 70% and 61 - 65% at a 1- and 14-day forecast horizon, respectively (supplementary Figure S9, second row).

There was little difference between calibration of the forecasts using the different catchment area definitions, especially compared to the difference between nominal and empirical coverage. The COVID-19 definition was the definition with the smallest overestimate of uncertainty for nominal coverage of 20% (Figure S9, first row), but this difference disappeared at higher nominal coverage values. There was also no difference between calibration at spatial scales (upper- vs. lower-tier local authority; Figure S9, first and second columns) or for future observed vs. future forecast cases (Figure S9, first and third column).

#### Probabilistic forecast accuracy

By forecast horizon, the forecasts made using the marginal distribution definition are consistently the least accurate (highest rWIS values); on the other hand the “nearby” heuristic and COVID-19 data definitions were generally the most accurate, with the other definitions (“nearest” heuristic, and emergency and elective data) falling in the mid-ranks (Figure 3A). There was only a small difference between definitions’ absolute rWIS values, which could suggest only small differences in probabilistic forecast accuracy, or no consistent trend in performance across forecast dates and/or locations. Finally, the average accuracy of forecasts made with the emergency and elective data definitions were very similar at all forecast horizons.

**Figure 3:**
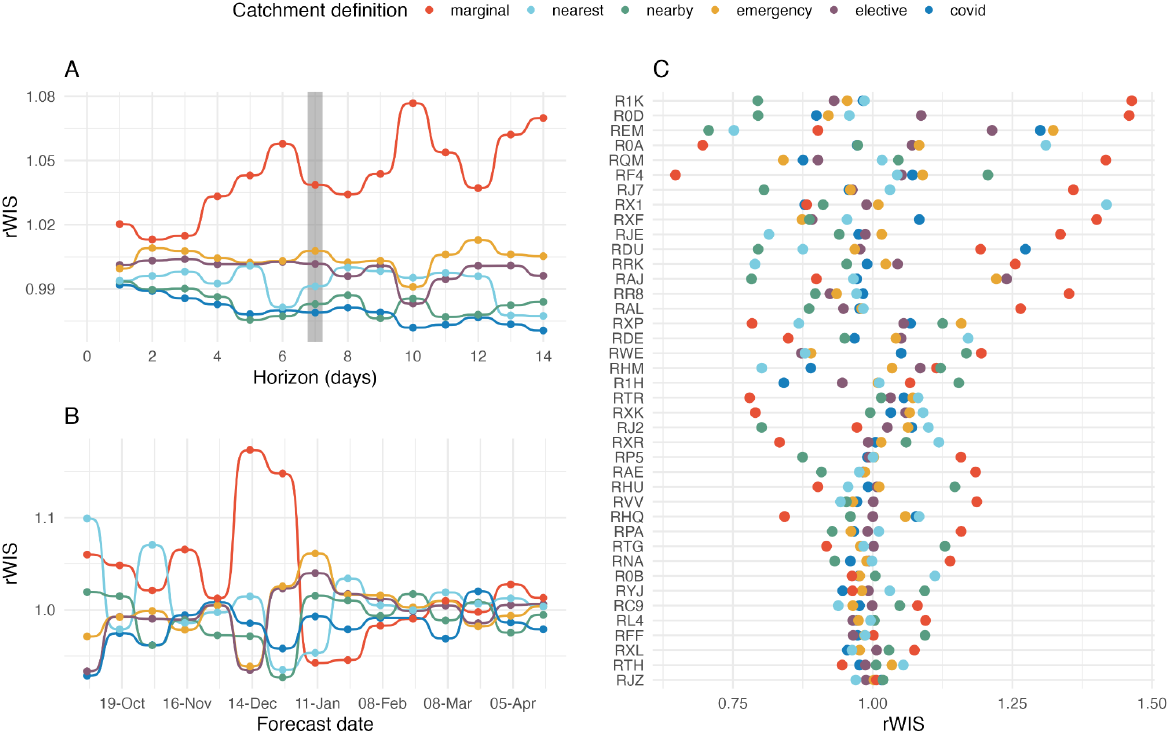
Forecasting performance under different hospital catchment area definitions (by UTLA) using future observed cases. (A) Median interval score (taken over all forecast dates and locations) for each forecast horizon, with the values highlighted for a 7-day forecast horizon. (B) Median interval score for each forecast date; 7-day forecast horizon. (C) Median interval score for the top 40 acute NHS Trusts (by total COVID-19 admissions); 7-day forecast horizon.

There was no clear best catchment area definition when we evaluated forecasts by forecast date (Figure 3B), with almost all definitions being either first- and last-ranked by rWIS for at least one forecast date (only the nearest heuristic was never first-ranked, and the elective admissions data definition was never last-ranked). The COVID-19 admissions data definition was first- or second-ranked by rWIS for the majority (9/15) of forecast dates at both a 7- and 14-day forecast horizon (supplementary Figure S10). At the same time, the definition was only last-ranked once at a 7-day horizon, and was never last-ranked at a 14-day horizon. The “nearby” heuristic performed comparably to the COVID-19 definition, and especially at a 7-day forecast horizon there was little to differentiate them. Forecasts made with the marginal distribution definition had the most variable accuracy, but were especially poor during December 2020. As noted previously, it was during this period that local COVID-19 case trends were more heterogeneous as a result of the emergence of Alpha and local social distancing regulations. These results therefore suggest that it is more important to use a local catchment area definition where there is heterogeneity in local case trends. Finally, we saw again that the emergency and elective admissions data definitions performed similarly across all forecast dates.

Again, there was no clear best definition when we evaluated forecasts by location, but there was more variation between definitions (Figure 3C). The COVID-19 definition was the first- or second-ranked definition more frequently than other definitions: (approx. 45% and 50% for a 7- and 14-day horizon, respectively; supplementary Figure S10). Again, the “nearby” heuristic also performed well (first- or second-ranked for approximately 35% and 40% of Trusts for a 7- and 14-day forecast horizon, respectively). However, the COVID-19 was more consistent than the nearby heuristic: while the COVID-19 definition was last-ranked for only 5% of Trusts, the nearby heuristic was last-ranked for 20%. Interestingly, the marginal distribution baseline definition was first-ranked, that is, it resulted in more accurate forecasts than other definitions for approximately 30% of locations, yet it also resulted in the least accurate forecasts in approximately 40% of locations (supplementary Figure S10B).

We found no change in relative forecast accuracy for any of the catchment area definitions when using LTLA-level catchment area definitions compared to UTLA-level definitions (Figure S11). Forecasts either performed comparatively or there was no clear pattern to differences when forecasts were evaluated by forecast horizon (Figure S11A), forecast date (Figure S11B) or location (Figure S11C).

When using forecasts of future cases instead of the retrospectively known case trajectories to make forecasts, the choice of catchment area definition had the biggest effect on probabilistic forecast accuracy at a 14-day forecast horizon (Figure 4). The marginal distribution definition had the largest rWIS value (rWIS = 1.53; Figure 4A). This poor performance was linked to a few forecast dates (4 and 18 October, 29 November and 13 December 2020; Figure 4B); for forecast dates in January 2021 onwards, the relative accuracy of all definitions was comparable. The nearest hospital heuristic resulted in the most accurate forecasts for a 14-day horizon (rWIS = 0.84; Figure 4A), largely due to particularly good relative forecast performance on 13 and 27 December 2020 (Figure 4B). All other definitions had rWIS values in the range 0.92 - 0.96 (Figure 4A). At shorter forecast horizons, the relative performance of all definitions was comparable, although the marginal distribution definition was consistently one of the worst-performing definitions.

**Figure 4:**
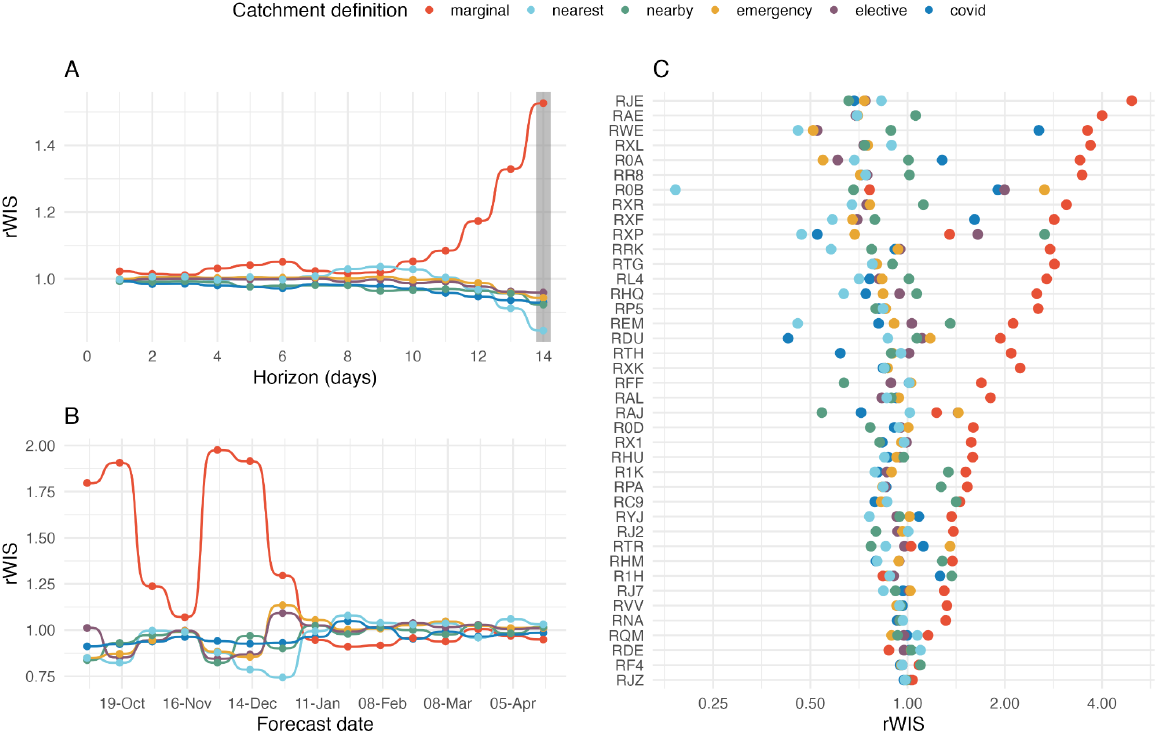
Forecasting performance under different hospital catchment area definitions using future forecast cases. (A) Median interval score (taken over all forecast dates and locations) for each forecast horizon, with the values highlighted for a 14-day forecast horizon. (B) Median interval score for each forecast date; 14-day forecast horizon. (C) Median interval score for the top 40 acute NHS Trusts (by total COVID-19 admissions); 14-day forecast horizon.

When considering the rWIS rankings by forecast date and location, the COVID-19 data definition stood out (Figure S11): it was first- or second-ranked for 7/14 forecast dates for both a 7- and 14-day forecast horizon, and for approximately 35% and 50% of locations for a 7- and 14-day forecast horizon. Although the nearby hospitals heuristic performed comparably to the COVID-19 definition as measured by top rWIS rankings, it was less consistent when evaluated by location: it was ranked in the bottom-two for 40% of locations, compared to only 21% and 12% for the COVID-19 definition for 7- and 14-day horizons, respectively.

## Discussion

This study systematically evaluated the impact of the choice of hospital catchment area definitions on the accuracy of local-level forecasts of COVID-19 hospital admissions during September 2020 - April 2021. While previous studies have set out to define hospital catchment areas or catchment populations [4–8,14], or to quantitatively evaluate local-level hospital admissions forecasts [1], none have considered the sensitivity of admissions forecasts to the choice of catchment area definitions.

At longer forecast horizons, the small improvement in forecast accuracy acquired from using any sensible catchment area definition is overshadowed by the decline in performance when uncertainty and/or inaccuracy of future cases is included. As noted previously [1], the benefit of including leading predictors, such as cases, is limited by the quality of forecasts of these predictors.

Our study focussed on only a single forecasting model, namely a scaled convolution of local cases and a delay distribution. We showed previously this model makes accurate forecasts of local-level COVID-19 hospital admissions, under the condition that we can make consistently good case forecasts [1]. As our aim here was to evaluate the relative performance of the catchment area definitions, this condition is not a limitation. However, we note that other forecasting models, including ensemble forecasts, may produce better forecasts in some scenarios, especially if these forecasts are made in real time.

Overall, we found that the choice of hospital catchment area definition affected the accuracy of hospital admission forecasts. The marginal distribution definition consistently resulted in the least accurate forecasts: it was clearly important to apply some definition of catchment areas. Moreover, forecasts made using a context-specific catchment area definition based on COVID-19 admissions data were the most accurate (or one of the most accurate) amongst all non-trivial catchment area definitions: the COVID-19 admissions data definition outperformed the other definitions when evaluated by forecast horizon and was highly-ranked more often when evaluated by forecast date or location. In addition, the relative performance of this definition was less variable across forecast dates and locations. The nearby heuristic also performed well, but was markedly less consistent than the COVID-19 definition when forecasts were evaluated by location. In total, these results demonstrate clear value in understanding the factors that drive local admissions patterns and, subsequently, constructing catchment area definitions that are relevant to any given pathogen or health emergency.

The difference in performance between the COVID-19 admissions definition and simpler heuristics was small; however, we posit that there are settings for which the choice of catchment area definition may be more important. Here, we showed that COVID-19 cases in local authorities within a catchment area for NHS Trusts in England were often strongly correlated with each other. Where the trend in cases is broadly similar within a catchment area the exact catchment area weights will have little-to-no impact on forecasting performance. On the other hand, we would expect to see more of an effect where local case trends are more heterogeneous. We saw some evidence of this during December 2020, where local case trends were more heterogeneous and the marginal distribution baseline performed especially poorly.

This opens a number of options for future work. First, we can test our assertion that the choice of catchment area definition is more important in heterogeneous settings. We propose the best approach would be to develop a small scale simulation study, where the correlation between cases within a catchment area is systematically varied. This synthetic study would be the best-case scenario for seeing difference in forecast performance by catchment area definition. We should also determine for which diseases, if any, it is feasible that this heterogeneity could arise. At the same time, we can replicate the study framework for COVID-19 in other countries, or for other diseases entirely. Determining when and where context-specific definitions are less important is valuable in real-time outbreak response settings: instead of dedicating significant time to constructing disease- and country-specific catchment area definitions, it may be sufficient to use simple heuristics to determine hospital catchment areas. When data and/or people-hours are limited, valuable resources can then be directed at more urgent analyses.

We also note a number of other limitations to this study. First, we assumed that catchment areas were fixed over time. Although preliminary analyses of COVID-19 admissions in England did not show any substantial change in catchment areas over time, this assumption will not always hold and a time-varying definition may be more appropriate in other contexts. For example, if hospitals regularly reach 100% capacity, then patients may be transferred to other facilities, or if new hospitals are opened or old hospitals closed. Second, we only defined catchment areas up to lower-tier local authorities. In some scenarios, this could mask fine-scale variation in reported cases; however, as we saw no improvement when using lower- vs. upper-tier local authority definitions here, we think it is unlikely yet smaller subnational regions would make a difference here. Additionally, using a definition based on smaller subnational regions would require substantially higher computational resources to make the case forecasts, which was already limited by this requirement to upper-tier local authorities only.

## Conclusions

Using catchment area definitions derived from context-specific data can improve local-level hospital admissions forecasts. Where context-specific data is not available, using catchment areas defined by carefully-chosen heuristics are a sufficiently-good substitute. There is clear value in understanding what drives local admissions patterns, and further research is needed to understand the impact of different catchment area definitions on forecast performance where case trends are more heterogeneous.

## Data Availability

Fully reproducible code is available at https://github.com/epiforecasts/hospitalcatchment-forecast and https://github.com/epiforecasts/hospitalcatchment.utils.

## Declarations

### Ethics approval and consent to participate

Not applicable.

### Consent for publication

Not applicable.

### Availability of data and materials

All data used in this study are publicly-available. Daily Trust-level COVID-19 hospital admissions are published weekly by NHS England and were accessed via the *covid19.nhs.data* R package [29]. Daily case reports aggregated by UTLA are published daily on the UK Government dashboard and were accessed via the *covidregionaldata* R package [30]. The Trust-UTLA mapping is available in the R package *covid19.nhs.data* [29].

### Authors contributions

SM designed and led the study, wrote the analysis code, performed the analysis and interpreted the results, and drafted and edited the manuscript. SFunk contributed to the development of the study, to the interpretation of results, and read, revised and approved the final manuscript.

## Acknowledgements

We thank Dr. Sam Abbott for his useful contributions to early drafts of the manuscript.

