## Supplementary Information for "Quantifying the impact of hospital catchment area definitions on hospital admissions forecasts: COVID-19 in England, September 2020 - April 2021"

---

***Contents***

**1. Forecasting models** **2**
1.1. Rt case forecast 2
1.2. Case-convolution admissions forecast 2

**2. Forecast evaluation metrics** **2**

**3. Data exploration** **3**
3.1. COVID-19 cases and hospital admissions 3
3.2. Hospital catchment area definitions 5
3.2.1. Descriptive statistics 5
3.2.2. Quantitative comparison of catchment area definitions 7
3.2.3. Quantitative comparison of COVID-19 case time series 10

**4. Forecast evaluation** **12**
4.1. Calibration 12
4.2. Probabilistic forecast error 13
4.2.1. Retrospective forecasts 13
4.2.2. Real-time forecasts 14

---

### 1. Forecasting models

#### 1.1. Rt case forecast

Forecasts of COVID-19 cases by UTLA (n = 174) were made via estimates and forecasts of the time-varying effective reproduction number, $R_t$, whilst accounting for uncertainty in the delay distributions [20, 21]. A summary is given below.

UTLA-level cases at time $t$ were assumed to be generated from a negative binomial observation model with overdispersion $\phi$ and mean $D_t$ (representing the number of cases by date of report at time $t$) scaled by a day-of-the-week effect with an independent parameter for each day of the week ($\omega_{t \bmod 7}$). Cases by date of report are a convolution of the onset-to-report distribution ($\xi_\tau$) and cases by date of onset ($O_{t-\tau}$), which are themselves a convolution of infections ($I_{t-\tau}$) with the incubation period ($\xi_\tau$).

Infections at time $t$ are modelled as a convolution of previous infections with the generation interval distribution ($w_\tau$), scaled by $R_t$. Temporal variation in Rt is controlled by a Gaussian process ($GP$).

In mathematical notation:

$$C_t \sim NegBin(\omega_{t \bmod 7} D_t, \phi)$$

$$D_t = \sum_\tau \xi_\tau O_{t-\tau}$$

$$O_t = \sum_\tau \zeta_\tau I_{t-\tau}$$

$$R_t \sim R_{t-1} \times GP.$$

We assume a log-Normal incubation period with Normal-distributed hyperpriors on the mean ($\mu_\zeta \sim N(5.2, 1.1^2)$) and standard deviation ($\sigma_\zeta \sim N(1.52, 1.1^2)$) [23]. We assume a gamma-distributed generation time $w$ with Normal-distributed hyperpriors on the mean ($\mu_w \sim N(3.6, 0.7^2)$) and standard deviation ($\sigma_w \sim N(3.1, 0.8^2)$); this is derived from [22], but refit with the above log-Normal distributed incubation period [21]. We also assume a log-Normal onset-to-report delay distribution, again with Normal-distributed hyperpriors on the mean ($\mu_\xi \sim N(0.523, 0.101^2)$) and standard deviation ($\sigma_\xi \sim N(0.945, 0.083^2)$) [21]. We assume $\phi \sim 1/\sqrt{N(0,1)}$.

Forecasts of UTLA-level COVID-19 cases at time $t + h$ are then made by assuming that $R_{t+h} = R_t$, from which we estimated future infections $I_{t+h}$ and then cases $C_{t+h}$ by the model outlined above. Future changes in contact rates, mobility, or non-pharmaceutical interventions are not accounted for.

#### 1.2. Case-convolution admissions forecast

Trust admissions at time $t$ ($A_t$) are assumed to be generated from a negative binomial observation model with mean $\mu_t$ and overdispersion $\phi$. The mean is a convolution of estimated Trust-level COVID-19 cases ($C^*_{t-\tau}$) with the confirmation-to-admission delay distribution ($\xi_\tau$), then scaled by the Trust-level case-hospitalisation ratio ($\alpha$). In mathematical notation:

$$A_t \sim NegBin(\mu_t, \phi)$$

$$\mu_t = \alpha \sum_{\tau=0}^{30} \xi_\tau C^*_{t-\tau} .$$

The parameters of the model were estimated during model fitting with the following weakly informed priors:

$$\alpha \sim N(0.2, 0.1),$$
$$\xi \sim LogNormal(\mu_\xi, \sigma_\xi),$$
$$\mu_\xi \sim N(2.5, 0.5),$$
$$\sigma_\xi \sim N(0.47, 0.25),$$
$$\phi \sim 1/\sqrt{N(0,1)}.$$

We do not include any seasonal effect, as previous work (unpublished) showed that incorrectly including a day-of-the-week effect in the case-convolution model (implemented as a simplex with a Dirichlet prior) led to additional uncertainty and subsequently worse model performance when a day of the week effect could not be identified.

The case-convolution model is fit and forecasts made independently for each date-location pair. Six weeks of data are used for fitting, including a 2-week burn-in period.

### 2. Forecast evaluation metrics

**Weighted interval score.** The weighted interval score (WIS) comprises a weighted sum of interval scores for quantile forecasts of increasing widths; in this way, the full forecast distribution is summarised in a single value.

The interval score [24] of the central 100(1-α)% predictive interval of forecast *F* is given by

$$IS_\alpha(F, y) = (u_\alpha - l_\alpha) + \frac{2}{\alpha}(l_\alpha - y)1_{\{y < l_\alpha\}} + \frac{2}{\alpha}(y - u_\alpha)1_{\{y > u_\alpha\}},$$

where $l_\alpha$ and $u_\alpha$ are the lower and upper bounds of the central 100*(1-α)% interval forecast, $y$ is the true observed value, and $1_{\{\cdot\}}$ is the indicator function (equal to 1 when the expression inside is true, and 0 otherwise). The first term measures sharpness, and penalises wider interval forecasts; the second term penalises forecasts for overprediction (if the true value, $y$, lies below the lower bound $l_\alpha$); finally, the third term penalises for underprediction.

Given the point forecast and $K$ interval forecasts of width $1 - \alpha_k$, $k = 1,\ldots, K$, the WIS is then calculated as

$$WIS_{\alpha_{0:K}}(F, y) = \frac{1}{K+0.5} \left( w_0 |y - m| + \sum_{k=1}^{K} w_k IS_{\alpha_k}(F, y) \right),$$

where the standard choice is $w_0 = 1/2$ and $w_k = \frac{\alpha_k}{2}$ for $k = 1,..., K$ [24]. In our evaluation, we used $K = 2$ and $\alpha_1 = 0.5$, $\alpha_2 = 0.1$, corresponding to the central 50% and 90% prediction intervals, respectively.

**Relative weighted interval score.** The relative weighted interval score (rWIS) is an adjusted WIS value that allows us to compare the performance of different catchment area definitions over forecast dates or between Trusts, both of which vary in the number of admissions. The rWIS is defined as follows.

First, the pairwise-relative WIS, $\theta_{A,B}$, for models (or, for this study, catchment area definitions) $A$ and $B$ is defined as

$$\theta_{A,B} = (mean\, WIS\, of\, model\, A) \,/\, (mean\, WIS\, of\, model\, B),$$

where the mean WIS is the mean in the scenario of interest (e.g. to evaluate models' overall performance at a 7-day horizon, the mean is taken over all forecast dates and Trusts).

The rWIS for model A, $\theta_A$, is then defined as the geometric mean of the pairwise-relative WIS $\theta_{A,B_i}$, $i = 1,..., M$, excluding the baseline model. If model A has a smaller relative WIS than model B, then forecasts generated by model A are better than those generated by model B.

### 3. Data exploration

#### 3.1. COVID-19 cases and hospital admissions

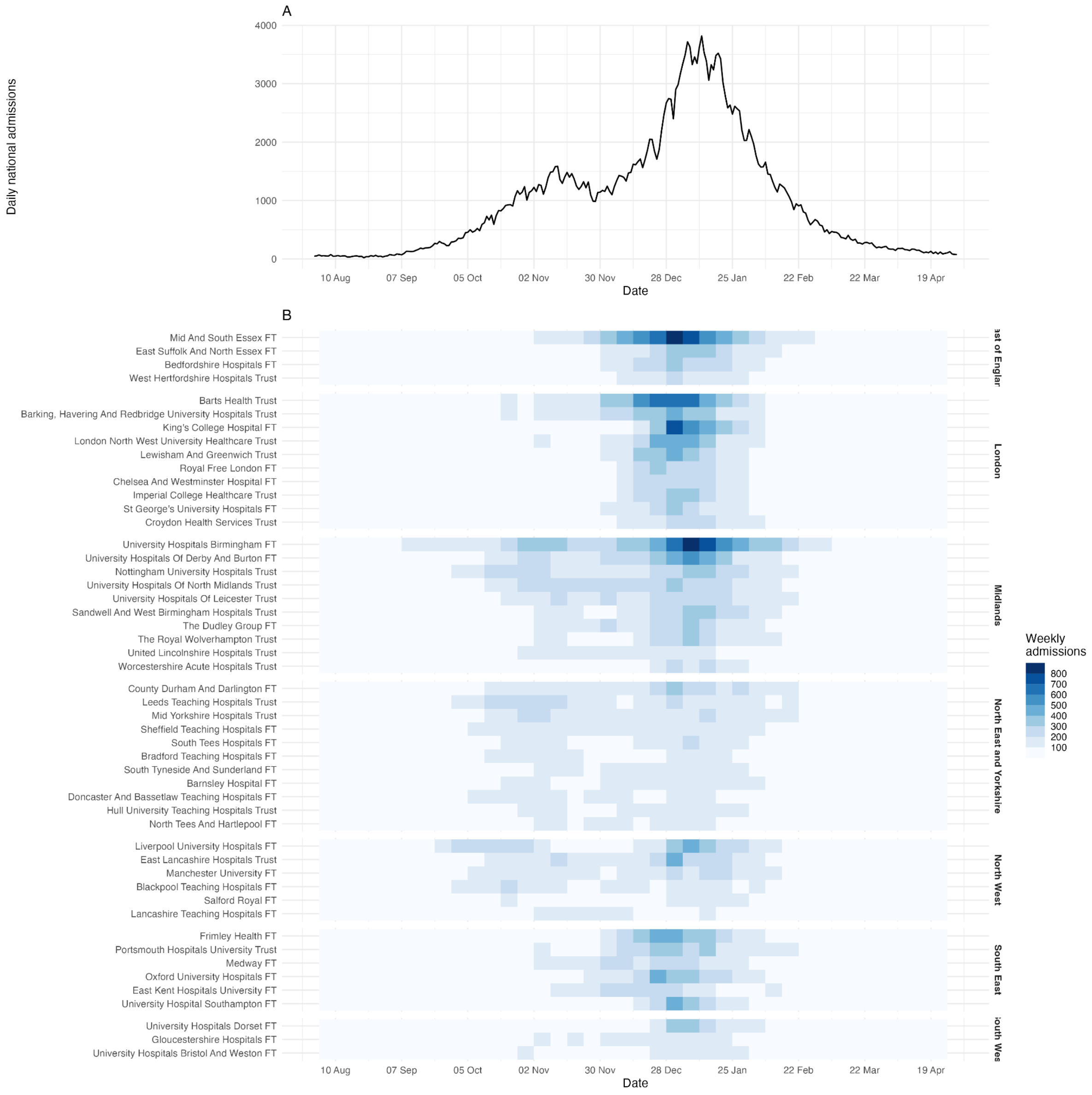


**Figure S1: Summary of COVID-19 hospital admissions August 2020 - May 2021.** (A) Daily COVID-19 hospital admissions across all acute NHS Trusts in England. (B) Weekly COVID-19 hospital admissions for the top-50 Trusts in England by total admissions August 2020 through April 2021. Trusts are grouped by NHS region, then ordered (top to bottom) by total admissions. “Foundation Trust” is abbreviated to “FT”.

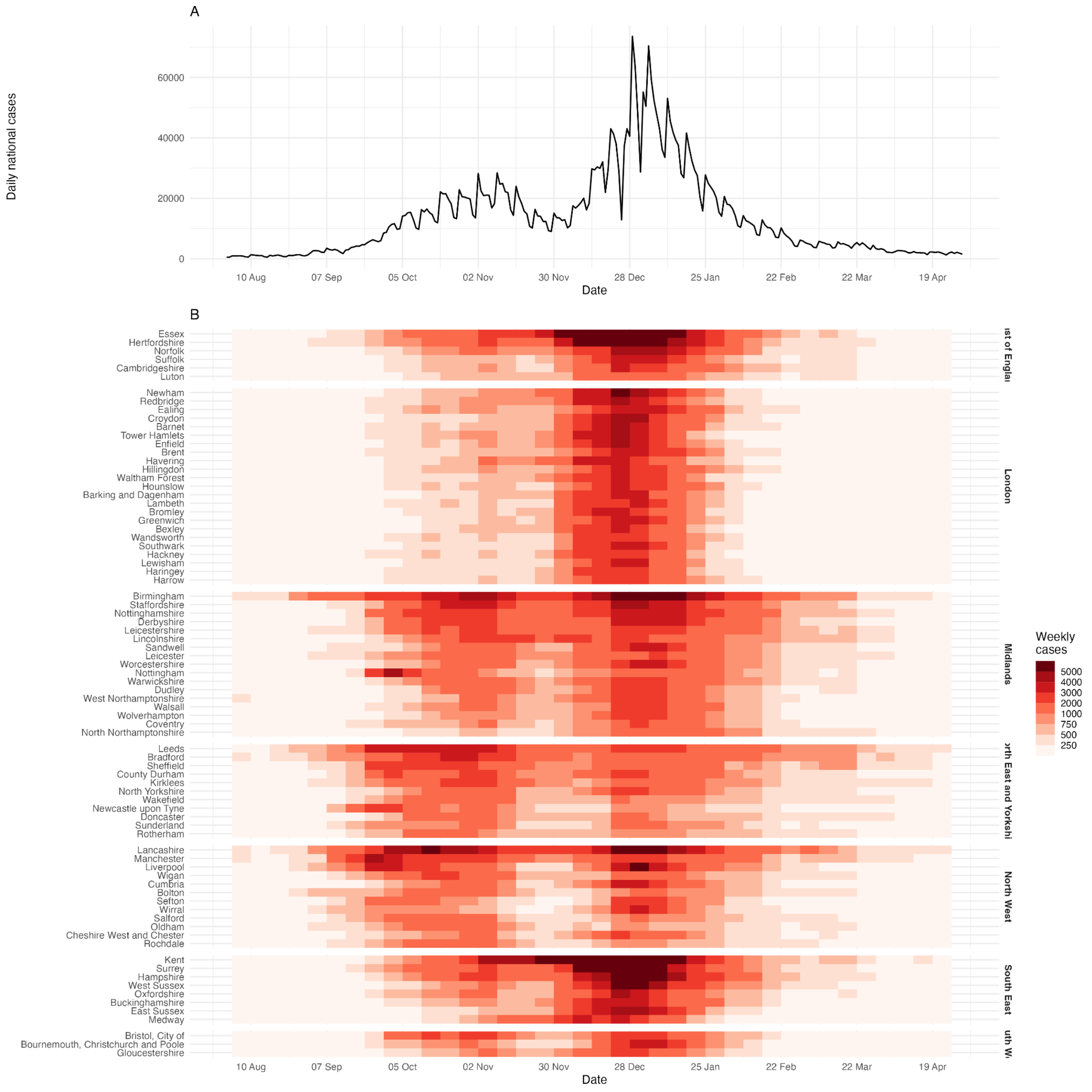


**Figure S2: Summary of COVID-19 cases August 2020 - May 2021.** (A) Daily COVID-19 cases in England. (B) Weekly COVID-19 cases for the top 80 upper-tier local authorities (UTLAs) in England by total cases August 2020 through April 2021. UTLAs are grouped by NHS region, then ordered (top to bottom) by total cases.

#### 3.2. Hospital catchment area definitions

##### 3.2.1. Descriptive statistics

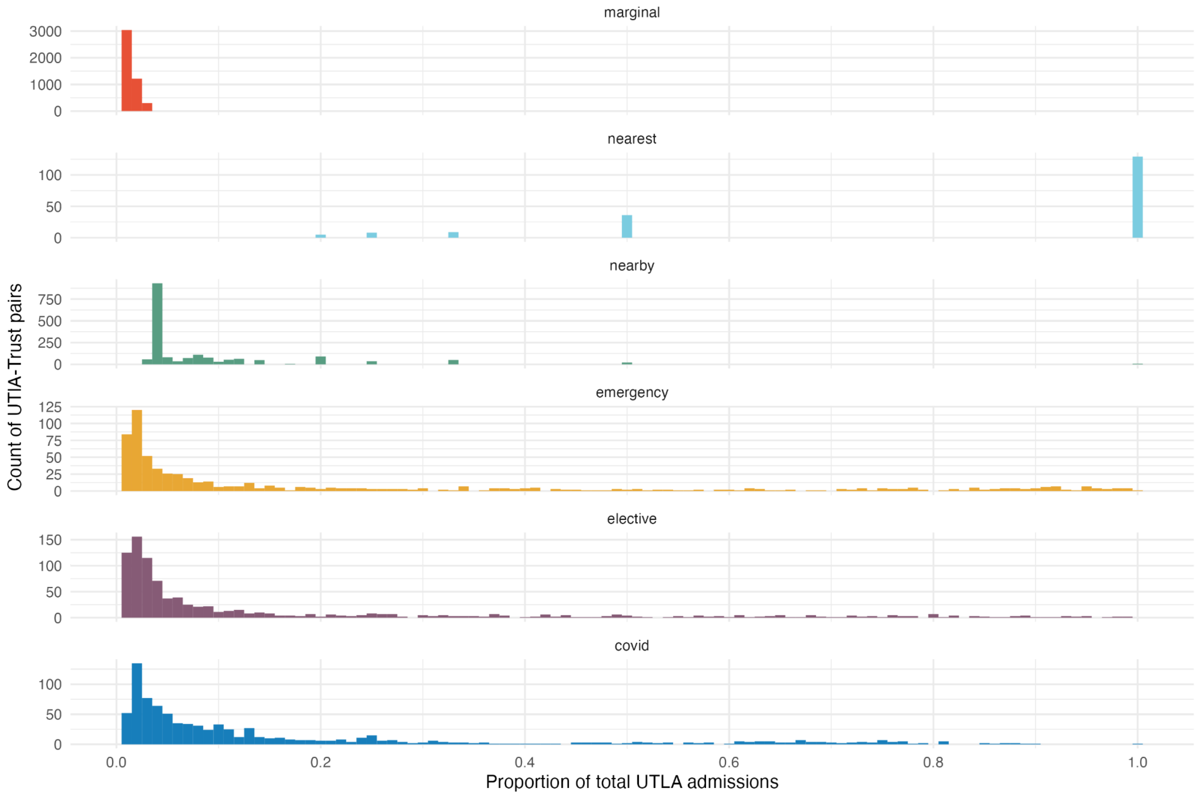


**Figure S3: Distribution of hospital catchment area weights.** For each upper-tier local authority (UTLA)-level catchment area definition, the panel shows the distribution of catchment area weights. Bins have width 0.01. Weights less than 0.01 (i.e. corresponding to less than 1% of total UTLA admissions) are not shown. Note that the y-axis scale is different for each catchment area definition, since each definition varies in the number of UTLA-Trust pairs with weights greater than or equal to 0.01.

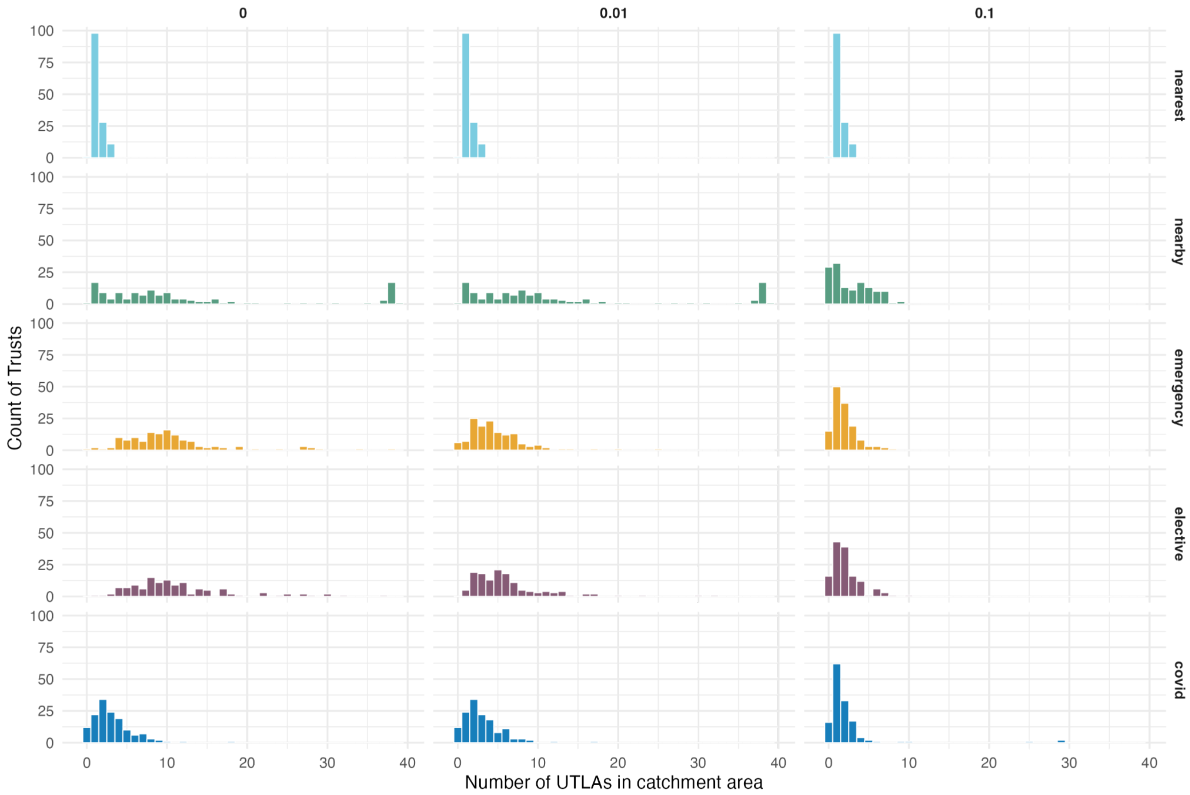


**Figure S4: Distribution of the number of local authorities in the catchment area definitions.** Distribution of number of upper-tier local authorities (UTLAs) in each Trusts catchment area from which at least x% are admitted to each Trust, where x = (left to right) 0%, 1%, 10% and 20%. The marginal distribution is not included as by definition each Trust includes all local authorities in its catchment area with weights greater than 0.01 but lower than 0.1, so the number of UTLAs in a catchment area will be 174 for all Trusts for x = 0, 1% and 0 for x = 10%.

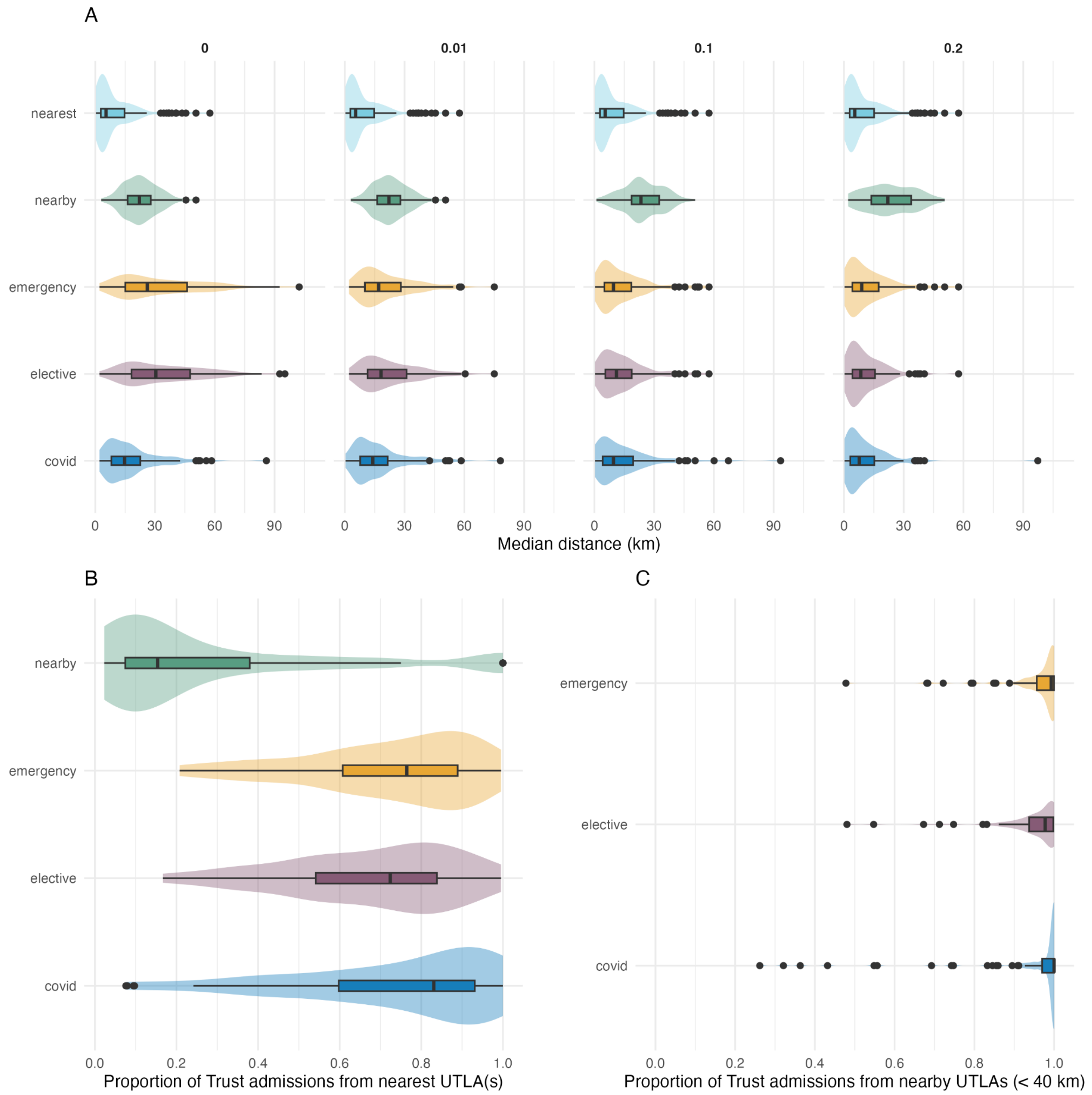


**Figure S5: Descriptive summary of hospital catchment area distances.** (A) Distribution of the median distance between a Trust and all local authorities in its catchment area from which at least x% are admitted to each Trust, where x = (left to right) 0%, 1%, 10% and 20%. The marginal distribution definition is now shown since the median distance will be the same for all Trusts. (B) Distribution of the proportion of a Trust's admissions that come from the nearest UTLA(s), as defined by the "nearest" catchment area definition, shown for the nearby, emergency, elective and COVID-19 catchment area definitions. (C) Distribution of the proportion of a Trust's admissions that come from any UTLA whose population centroid lies within a 40 km radius of the main Trust site, shown for the emergency, elective and COVID-19 catchment area definitions.

##### 3.2.2. Quantitative comparison of catchment area definitions

We defined the overlap-similarity metric to quantify the similarity between pairs of catchment area definitions for a given Trust. The *asymmetric overlap-similarity* between definitions $X$ and $Y$ relative to definition $X$ is defined as the proportion of the definition $X$ that is contained in the overlap with $Y$. The *overlap-similarity* between $X$ and $Y$ is then simply the minimum of the asymmetric overlap-similarity relative to each of $X$ and $Y$.

**Definition.** Let $X$ and $Y$ be two catchment area definitions, and let $x_{ij}$ and $y_{ij}$ be the proportion of admissions from local authority $i$ that go to Trust (hospital) $j$ for definitions $X$ and $Y$, respectively.

The asymmetric overlap-similarity between definitions $X$ and $Y$ relative to definition $X$ is

$$p_j^{(X,Y)} = \sum_i \min\{x_{ij}, y_{ij}\} / \sum_i x_{ij} \in [0, 1],$$

and the overlap-similarity between definitions $X$ and $Y$ is simply

$$p_j^{XY} = \min\{p_j^{(X,Y)}, p_j^{(Y,X)}\} \in [0, 1].$$

**Properties.**

a. The overlap-similarity is zero if, and only if, the two definitions do not both cover any of the same local authorities:

$$p_j^{XY} = 0 \Rightarrow \{x_{ij} = 0\} \vee \{y_{ij} = 0\} \forall i.$$

b. The asymmetric overlap-similarity relative to definition $X$ is equal to 1 if, and only if, the weights assigned by definition $X$ are less than or equal to the weights assigned by $Y$ for all local authorities:

$$p_j^{(X,Y)} = 1 \Leftrightarrow x_{ij} \leq y_{ij} \forall i.$$

c. The overlap-similarity metric is equal to 1 if, and only if, the two definitions assign exactly equal weights for all local authorities:

$$p_j^{XY} = 1 \Leftrightarrow x_{ij} = y_{ij} \forall i.$$

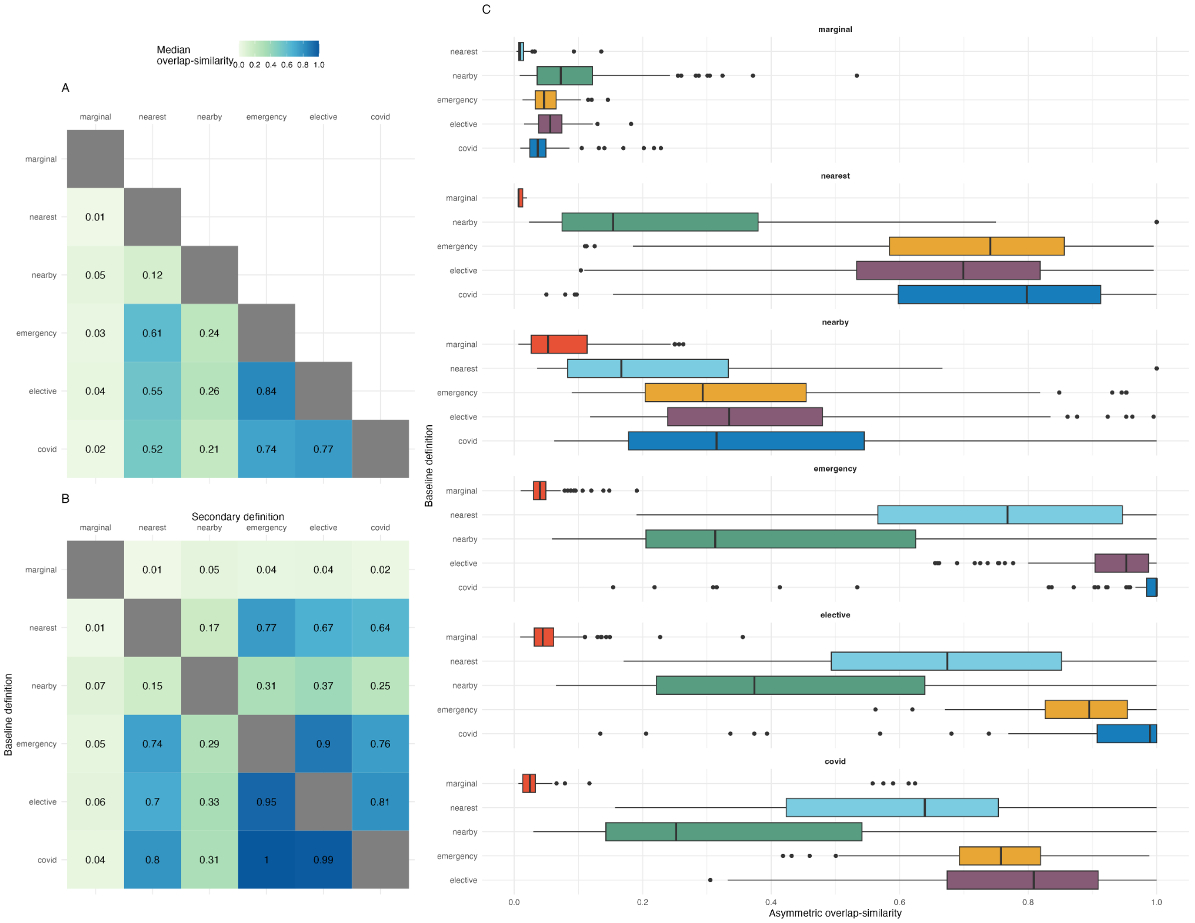


**Figure S6: A quantitative comparison of hospital catchment area definitions using the overlap-similarity metric.** (A) Median overlap-similarity value for each unordered pair of catchment area definitions across all Trusts, where the overlap-similarity is the minimum of the two asymmetric overlap-similarity values. (B) Median asymmetric overlap-similarity across all Trusts, relative to the baseline definition (y-axis). The values in each column are exactly the median values shown in the corresponding subpanel of C. (C) Distributions of asymmetric overlap-similarity values for each pair of catchment area definitions, relative to the baseline definition (y-axis).

##### 3.2.3. Quantitative comparison of COVID-19 case time series

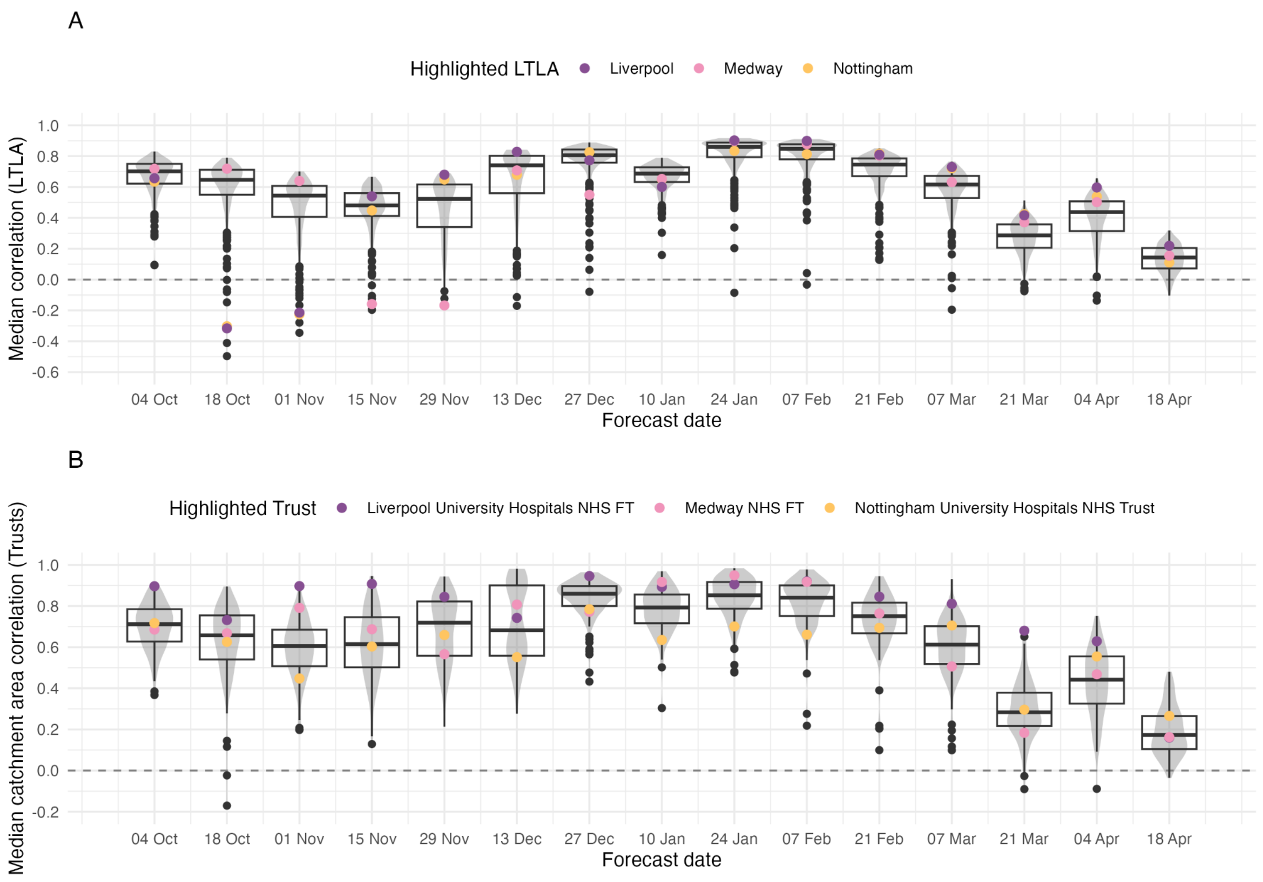


**Figure S7: Distribution of the median correlation between COVID-19 case time series by local authority.** For each forecast date and pair of lower-tier local authorities (LTLAs), we calculated the Pearson correlation between the time series of COVID-19 cases in each LTLA from 21 days before until 14 days after the forecast date. For each forecast date the distribution of the following are shown. (A) For each LTLA, the median correlation coefficient with all other local authorities. Three local authorities are highlighted: Liverpool (a city in North West England), Medway (a mostly rural local authority in South East England) and Nottingham (a city in the Midlands). (B) For each NHS Trust, the median correlation coefficient between all local authorities for which any catchment area definition assigns a weight of more than 10%. Three NHS Trusts are highlighted: Liverpool University Hospitals NHS Foundation Trust (local authorities Liverpool, Sefton, Knowsley, and West Lancashire), Medway NHS Foundation Trust (Medway and Swale), and Nottingham University Hospitals NHS Trust (12 local authorities including Nottingham, Charnwood, Ashfield and Mansfield).

### 4. Forecast evaluation

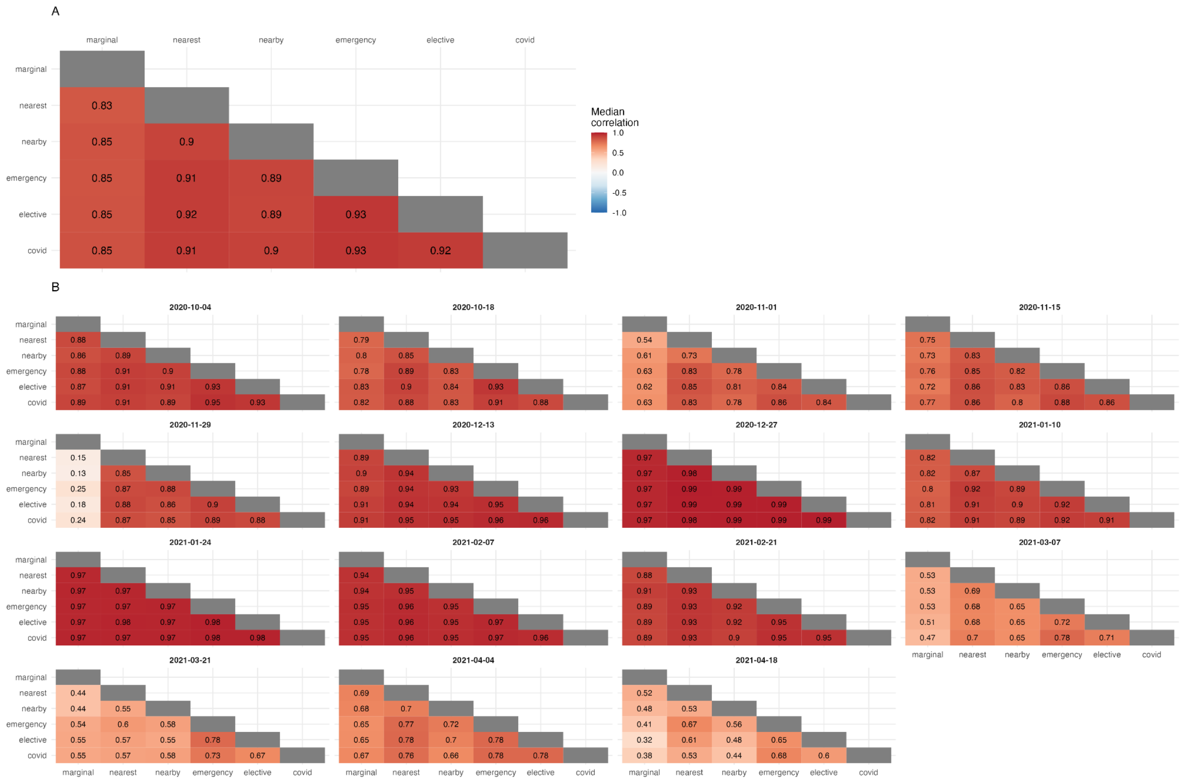


**Figure S8: Similarity of median hospital admissions forecasts made under six catchment area definitions.** (A) The median correlation coefficient: for each forecast date and location the Pearson correlation coefficient is calculated between the median forecasts for each pair of catchment area definitions. (B) The median correlation coefficient by forecast date: as above, but the median is calculated by forecast date.

### 4.1. Calibration

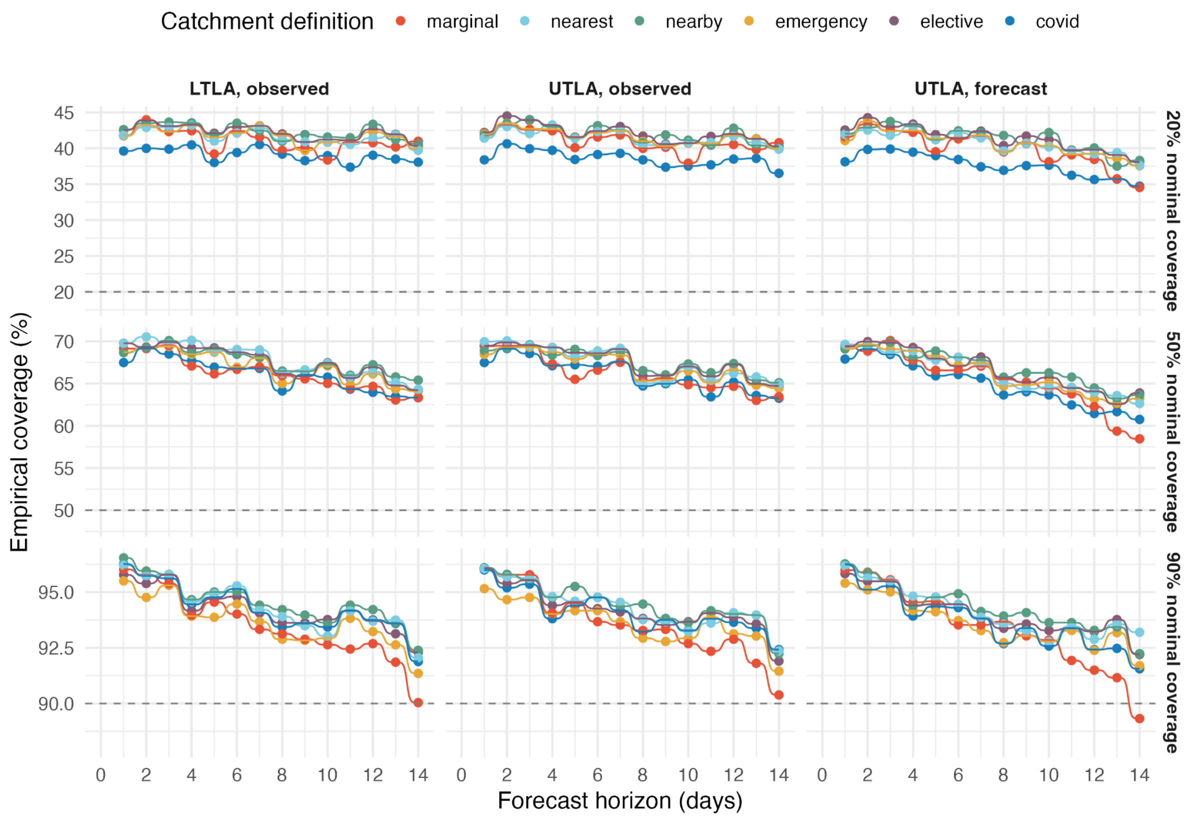


**Figure S9: Empirical coverage of forecasts.** Empirical coverage of forecasts made with each of the hospital catchment area definitions for nominal coverage of 20%, 50% and 90% (top-bottom); dashed horizontal line indicates nominal coverage, for reference. Scenarios are shown in columns: lower-tier local authority (LTLA) using future observed cases; upper-tier local authority (UTLA) using future observed cases; UTLA using future forecast cases.

### 4.2. Probabilistic forecast error

#### 4.2.1. Retrospective forecasts

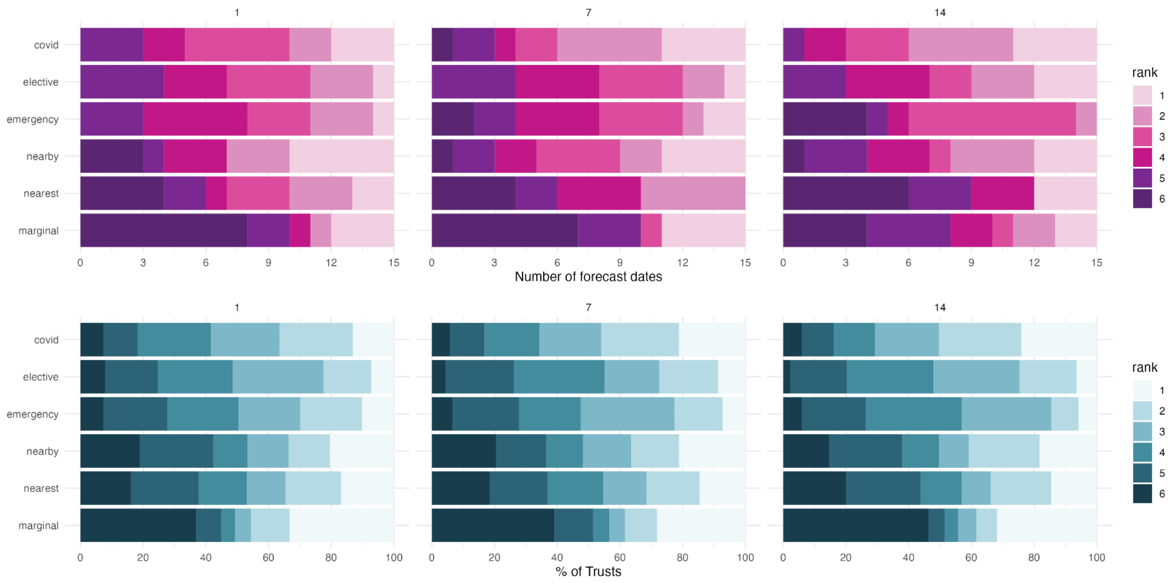

**Figure S10: Distribution of WIS rankings by forecast date and location for models using future observed cases.** Distribution of rankings (1 = best) of median WIS across (A) 15 forecast dates and (B) 138 acute NHS Trusts; for a forecast horizon of 1, 7 and 14 days.

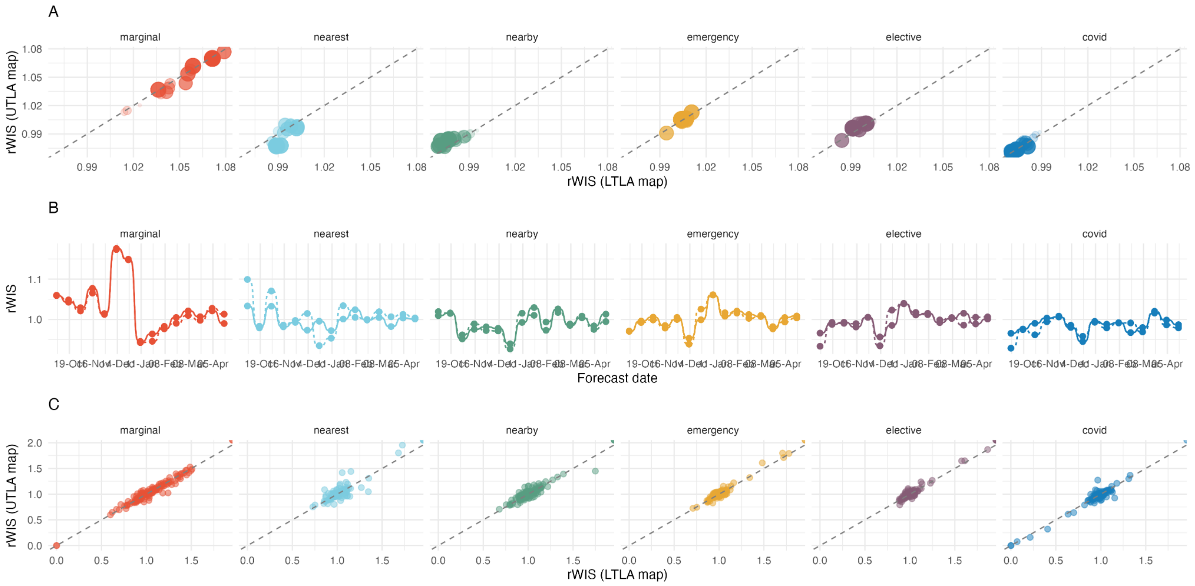


**Figure S11: Comparison of forecasting performance under different hospital catchment area definitions between LTLA and UTLA.** (A) By forecast horizon; increasing point size and opacity both denote increasing forecast horizon. (B) By forecast date, where the solid and dashed lines show the LTLA- and UTLA-level catchment area definitions, respectively. (C) By location (each point is an acute NHS Trust).

#### 4.2.2. Real-time forecasts

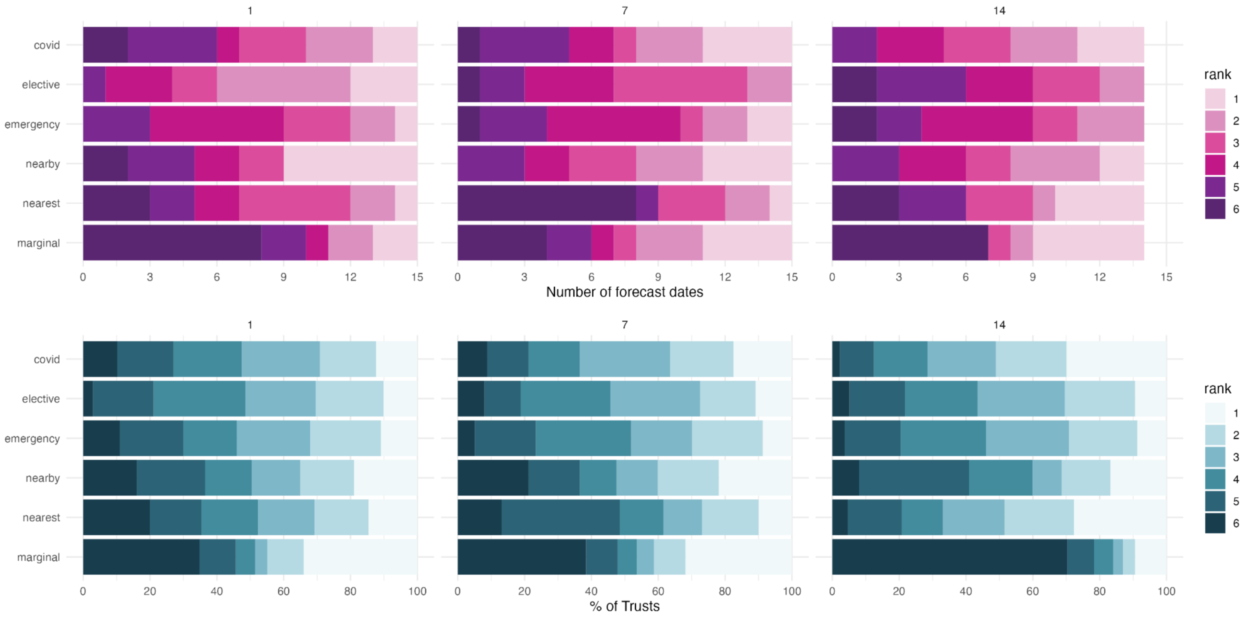


**Figure S12: Distribution of WIS rankings by forecast date and location for models using future forecast cases.** Distribution of rankings (1 = best) of median WIS across (A) 15 forecast dates and (B) 138 acute NHS Trusts; for a forecast horizon of 1, 7 and 14 days.
